# Hospital initiation of benzodiazepines and Z-drugs in older adults and discontinuation in primary care

**DOI:** 10.1101/2020.07.24.20161711

**Authors:** Seán Coll, Mary E Walsh, Tom Fahey, Frank Moriarty

## Abstract

**Objective:** To examine factors associated with continuation of hospital-initiated benzodiazepine receptor agonists (BZRAs) among adults aged ≥65 years, specifically instructions on hospital discharge summaries.

**Methods:** This retrospective cohort study involved anonymised electronic record data on prescribing and hospitalisations for 38,229 patients aged ≥65 from forty-four GP practices in Ireland 2011-2016. BZRA initiations were identified among patients with no BZRA prescription in the previous 12 months. Multivariate regression examined whether instructions on discharge messages for hospital-initiated BZRA prescriptions was associated with continuation after discharge in primary care and time to discontinuation.

**Results:** Most BZRA initiations occurred in primary care, however the rate of hospital-initiated BZRAs was higher. Almost 60% of 418 hospital initiations had some BZRA instructions (e.g. duration) on the discharge summary. Approximately 40% (n=166) were continued in primary care. Lower age, being prescribed a Z-drug or great number of medicines were associated with higher risk of continuation. Of those continued in primary care, in 98 cases (59.6%) the BZRA was discontinued during follow-up (after a mean 184 days). Presence of instructions was associated with higher likelihood of discontinuation (hazard ratio 1.67, 95%CI 1.09-2.55).

**Conclusions:** Improved communication to GPs after hospital discharge may be important in avoiding long-term BZRA use.

## Introduction

Benzodiazepines and z-drugs (i.e. benzodiazepine receptor agonists or BZRAs) are frequently prescribed medicines, most often being used in the management of insomnia and anxiety. As with any medicine, the benefits for these clinical indications must be balanced against the risk of harm for the individual. Risks associated with BZRAs are of particular concern when prescribed to older people and/or on a long-term basis as often these risks outweigh the benefits.^1^ Despite this, the prevalence of use of these agents is high internationally, in particular long-term use in older adults.^2^ In the United States, trends indicate that outpatient use of these medicines has increased significantly in recent decades.^3^ Although Ireland has among the highest levels of long-term use among older adults among OECD countries,^2^ prescribing rates of long-acting benzodiazepines have been decreasing slowly since 2005.^4^

Older people may be especially prone to adverse effects from long-term BZRA use as pharmacokinetic and pharmacodynamic changes in ageing can lead to increased retention time resulting in prolonged, potentiated effects.^5^ These factors can result in hangover effects, and contribute to increased risk of falls, fractures and road traffic accidents among others.^6^ This issue is highlighted in the American Geriatrics Society’s Beer’s criteria which states that they should generally be avoided due to increased risks of falls and delirium, but that they are suitable in some specific conditions.^7^ The European STOPP (Screening Tool for Older Persons’ Prescriptions) criteria deem use in older people for a period of greater than a month to be potentially inappropriate prescribing.^6^ These aformentioned sequelae of long term benzodiazepine and z-drugs prescribing can also increase healthcare costs consequently burdening others in the health system.^8^

In addition, long-term use can result in the development of tolerance and dependence, which presents an example of a barrier to reduce (i.e. deprescribe) these agents. A systematic review of barriers to deprescribing found that fear was the most common patient barrier for BZRAs, including withdrawal symptoms and condition deterioration.^9^ Prescriber barriers are also a significant factor to consider, for example inertia, fear of the unknown and incomplete information on the rationale for prescribing have been shown to prevent change.^10^ Older adults may receive care from several prescribers, where they have multiple conditions, and may have frequent hospital admissions. This can further impede deprescribing when a general practitioner (GP) is trying to optimise treatment in the context of transitions of care and medications initiated by multiple other prescribers. Therefore, improving communication of information from initiating prescribers across the secondary care-primary care interface may address some of these barriers to deprescribing. Studies have shown that providing relevant information in hospital discharge summaries encouraged faster discontinuation of proton pump inhibitors where long-term use is not indicated.^11,12^ Although previous research has found hospital admission are associated with increased initiation of benzodiazepines,^13–16^ the impact of hospital discharge summary information on deprescribing has not been considered to date.

Therefore this study aims to examine factors associated with continuation of hospital-initiated BZRAs among among adults aged 65 years and over, and specifically whether instructions on hospital discharge summaries are associated with their discontinuation.

## Methods

### Study design and participants

This is a retrospective cohort study using anonymised data from 44 GP practices in Ireland between 2011 and 2016. Data on prescriptions, consultations, demographics, and hospitalisations were extracted from the GP patient management software Socrates for patients aged 65 years and over. Patients were included in the present study if they were classified as BZRA naïve at any point during follow-up. This was defined as having received no prescription (either from their GP or at hospital discharge) for a BZRA (any benzodiazepine or Z-drug medication) in the previous 365 days. These were identifed in hospital discharge data based on brand and non-proprietary names for medications, including potential misspellings, and in the GP prescriptions data based on World Health Organisation Anatomical Theraputic Classification (ATC) codes (N05CF, N05CD, N05BA, and N03AE).

### Variables

BZRA initiations were identified in hospital or primary care among BZRA naïve patients. The following characteristics were described: patient age, sex, and number of regular medicines, type of BZRA initiated (benzodiazepine, Z-drug or both), hospital length of stay, and patient health cover. Number of regular medicines was defined as the number of unique fourth level ATC codes (i.e. chemical subgroup/drug class) prescribed by their GP in the 12 months preceding BZRA initiation, categorised as 0-4, 5-9, 10-14, or 15 or more medicines. Health cover was categorised as GMS scheme cover (providing a range of health services at low or no cost, including medication), Doctor Visit card (providing only GP visits free at the point of access) or neither of these (i.e. private patients). GMS scheme and Doctor Visit card eligibility are based on income means testing, and therefore eligible patients tend to be more socioeconomically deprived.^17^ For hospital-initiated BZRAs, all discharge messages (sent from the hospital to the patient’s GP) were reviewed manually to identify if they contained any instructions relating to the newly initiated BZRA, for example on tapering (e.g. decreasing doses or the words “weaning”, “reducing”, “tapering”, “decreasing”), duration (indicated by an amount of time, or a number of tablets), marked for review (e.g. “for review by GP”, “consider weaning off”) or as required use (e.g. “prn”, “as needed”).

We determined the proportion of hospital initiations that were continued in primary care (prescribed within 90 days of discharge). Of those that were continued, the time to discontinuation was calculated.We used the specified duration of each prescription, or where this was implausibly long (i.e. beyond the 6 month validity of prescriptions in Ireland) we inferred from the quantity and dosing of the specific drug. We then defined discontinuation as a BZRA-free period of ≥135 days after the duration of the latest prescription. This was chosen as a margin of 1.5 times the typical 90 day prescription duration.

### Analysis

Descriptive statistics were generated to characterise both primary care and hospital BZRA initiations. Rate of initiation in primary care was expressed as a function of the number of prescriptions issued to BZRA naïve patients (considering all items prescribed on a single date to a patient as one prescription), and the rate of initiation in hospital as a function of the number of hospitalisations for BZRA naïve patients. Rates were also expressed in terms of time spent naïve in primary care and hospital. For those hospital-initiated BZRAs, we examined factors associated with continuation in primary care using multivariable generalised linear regression models with Poisson distributions to generate relative risks with 95% confidence intervals (CIs). Among those BZRAs that were continued, we examined factors, in particular presence of instructions, associated with time to discontinuation using multivariable Cox regression to generate hazard ratios with 95% CIs. We accounted for potential clustering of repeated initiations in the same patient by estimating robust standard errors. STATA v14 was used for analyses and statistical significance was assumed at p<0.05.

## Results

Among 38,229 patient, there were 7,876 BZRA initiations during the study, and 7,458 of these were initiated in primary care and 418 were hospital-initiated. This represents 6.7 initiations per 1,000 prescriptions to BZRA naïve patients in primary care, and 25.8 initiations per 1,000 hospitalisations involving BZRA naïve patients. In terms of time exposed, there were 0.29 initiations per 1,000 days naïve in primary care, and 1.90 per 1,000 days naïve during hospital stays. Hospital initiations were more often to older patients, men, and to those with GMS or no health cover (Table 1). For hospital-initiated BZRAs, 59.1% of these had some level of instructions on the discharge message. Most often this was specifying use as required, a duration of prescription, or instructions to taper or review.

**Table 1.** Descriptive statistics relating to primary care and hospital initiation of BZRAs.

|  | **Initiations** |  |
| --- | --- | --- |
|  | **Primary Care**<br>**n=7,458** | **Hospital**<br>**n=418** |
| **Age, mean (SD)** | 75.9 (8.0) | 79.0 (8.3) |
| **Male, n (%)** | 2,784 (37.3) | 204 (48.8) |
| **BZRA type, n (%)** |  |  |
| Benzodiazepine | 3,966 (53.2) | 209 (50.0) |
| Z-drug | 3,339 (44.8) | 187 (44.7) |
| Both | 153 (2.1) | 22 (5.2) |
| **Health cover, n (%)** |  |  |
| GMS scheme | 5,132 (68.8) | 298 (71.3) |
| Doctor visit card | 896 (12.0) | 21 (5.0) |
| Private patient | 1,430 (19.2) | 99 (23.7) |
| **Number of medicines, mean (SD)** | 9.3 (6.1) | 10.0 (6.9) |

BZRA: benzodiazepine receptor agonist, GMS: General Medical Services, SD: standard deviation.

In 39.7% of cases of hospital inititation (n=166/418) there was a BZRA prescription in primary care within 90 days of discharge. Factors associated with continuation following discharge included lower age, being initiated on a Z-drug (compared to a benzodiazepine) or being on a more regular medicines (Table 2). Of these 166 cases, for 98 (59.6%) the BZRA was deprescribed during follow-up (mean time to discontinuation was 184 days (SD 230), Figure 1). Presence of instructions on the hospital discharge summary was independently associated with likelihood of discontinuation (adjusted hazard ratio 1.67, 95% CI 1.09 to 2.55, Table 3).

**Table 2.** Factors associated with BZRA continuation in primary care following hospital initiation (n=418)

| **Covariates** | **Adjusted relative risk (95% confidence interval)** | **P value** |
| --- | --- | --- |
| **Instructions** | 0.93 (0.74 to 1.16) | 0.518 |
| **Age (years)** | 0.97 (0.95 to 0.98) | <0.001 |
| **BZRA type (ref benzodiazepine)** |  |  |
| Z-drug | 1.35 (1.08 to 1.70) | 0.009 |
| Both | 1.17 (0.65 to 2.11) | 0.603 |
| **Health cover (ref GMS scheme)** |  |  |
| Doctor visit card | 0.63 (0.33 to 1.23) | 0.177 |
| Private patient | 0.80 (0.59 to 1.08) | 0.142 |
| **Male** | 0.96 (0.77 to 1.21) | 0.740 |
| **Number of medicines (ref 0-4)** |  |  |
| 5-9 medicines | 1.63 (1.04 to 2.55) | 0.033 |
| 10-14 medicines | 2.00 (1.30 to 3.08) | 0.002 |
| 15 or more medicines | 2.24 (1.48 to 3.41) | <0.001 |
| **Length of stay (ref 1st quartile)** |  |  |
| 2nd quartile | 1.20 (0.87 to 1.67) | 0.269 |
| 3rd quartile | 1.18 (0.88 to 1.58) | 0.273 |
| 4th quartile | 0.74 (0.50 to 1.08) | 0.121 |
| Missing | 0.63 (0.38 to 1.05) | 0.076 |

BZRA: benzodiazepine receptor agonist, GMS: General Medical Services.

**Table 3.** Factors associated with time to BZRA discontinuation among those hospital initiations that were continued in primary care (n=166)

| **Covariates** | **Adjusted hazard ratio**<br>**(95% CI)** | **P value** |
| --- | --- | --- |
| **Instructions** | 1.67 (1.09 to 2.55) | 0.018 |
| **Age (years)** | 0.97 (0.95 to 1.00) | 0.061 |
| **BZRA type (ref benzodiazepine)** |  |  |
| Z-drug | 0.89 (0.56 to 1.42) | 0.624 |
| Both | 0.66 (0.17 to 2.54) | 0.551 |
| **Health cover (ref GMS scheme)** |  |  |
| Doctor visit card | 0.68 (0.17 to 2.77) | 0.592 |
| Private patient | 1.51 (0.97 to 2.36) | 0.067 |
| **Male** | 0.88 (0.56 to 1.37) | 0.566 |
| **Number of medicines (ref 0-4)** |  |  |
| 5-9 medicines | 1.77 (0.80 to 3.92) | 0.156 |
| 10-14 medicines | 1.59 (0.75 to 3.39) | 0.225 |
| 15 or more medicines | 1.77 (0.87 to 3.59) | 0.114 |
| **Length of stay (ref 1st quartile)** |  |  |
| 2nd quartile | 0.77 (0.40 to 1.50) | 0.441 |
| 3rd quartile | 0.69 (0.37 to 1.31) | 0.256 |
| 4th quartile | 0.60 (0.28 to 1.28) | 0.183 |
| Missing | 0.99 (0.53 to 1.87) | 0.985 |

BZRA: benzodiazepine receptor agonist, GMS: General Medical Services.

**Figure 1.**
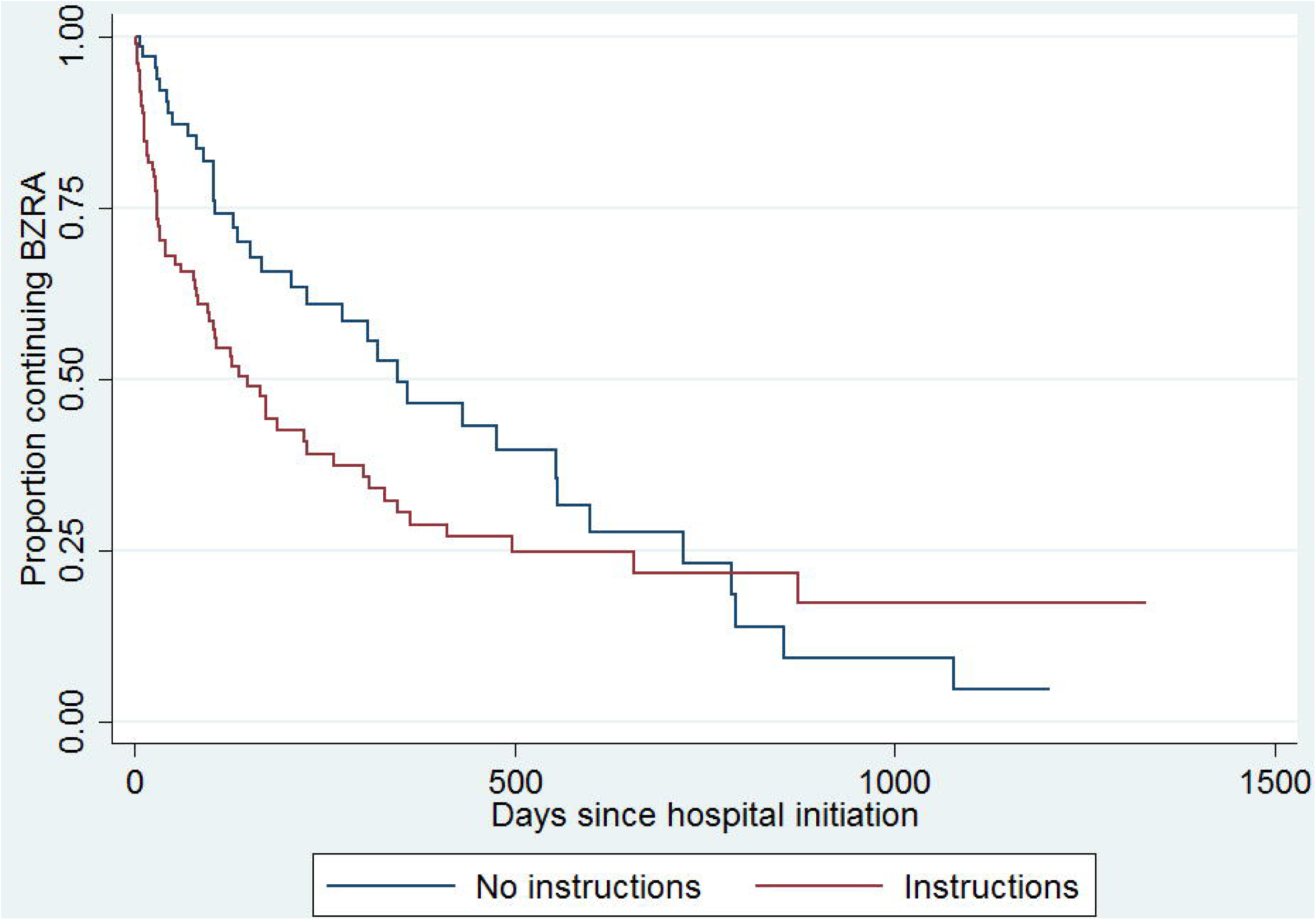
Kaplan Meier curve of time to benzodiazepine receptor agonist discontinuation among hospital initiations with and without instructions

## Discussion

Our study shows that inclusion of instructions on hospital discharge summaries is associated with shorter time to discontinuation of BZRAs amongst older people in primary care. Previous studies on BZRAs being prescribed in hospital have identified specific cases of potentially inappropriate prescribing^13^, found that the period 3 months before and after hospitalisation is associated with increased rates of initiation of benzodiazepines^14^, and assessed the influence of specific in-patient departments on use.^15^ Another study found increased rates of BZRA initiation and continuation in older adults specifically.^16^ The positive association of number of medicines or polypharmacy with continuation is consistent with previous research on benzodiazepines initated after hospitalisation.^14,15,16^ Those started on a z-drug alone rather than a benzodiazepine were more likely to be continued following discharge, possibly reflecting a different perception of these medicines in the eyes of prescribers. Although health cover did not show a significant association in our study, previous evidence suggests an inverse relationship between socioeconomic status and BZRA prescribing rates.^18^

A strength of this study is the use of actual hospital prescribing at discharge, compared to previous studies using GP prescribing after discharge only. Our study is limited by a lack of information on use of non-prescription or non-pharmacological treatments, such as cognitive behavioural therapy (CBT), which may influence rates of prescribing. We also were unable to account for prescribing indication for BZRAs, including scenarios such as palliative care where long-term prescription may be appropriate. Although information on co-morbidities was lacking, we adjusted for prescribed medicines as a proxy for illness burden. For our primary analysis, we would not expect co-morbidities to differ systematically between discharge messages with or without instructions.

However our study examines a novel question and suggests that when additional instructions relating to hospital-initiated BZRAs are conveyed to GPs, earlier discontinuation of these medications can be facilitated. This sharing of information has been identified as an essential component of clinical handovers^19^ and hospital discharge summaries are a conduit for this information at secondary-primary care interfaces. The literature has identified that amongst hospital-based junior doctors and community-based GPs particularly, there can be a disconnect in intraprofessional perceptions of what to include on a hospital discharge summary^20^, and that more training and inclusion in medical curriculums may improve continuity of care.^20,21^

Our findings are consistent with systematic review evidence that identifies lack of information on discharge summaries as a patient safety risk.^22^ Without instructions from the original prescriber, GPs may be reluctant to reduce or stop a BZRA and thus short-term prescriptions can develop into long-term use. Systems for communication between prescribers at transitions of care, such as shared electronic medical records or electronic discharge summaries, should incorporate structured fields to record important information to support medication review and deprescribing, including indication, intended duration of use and review timeframe. IT-based summaries in particular have been shown to be preferable to both patients and prescribers, because of their structured and accessible nature.^23^

Long-term BZRA use may also be avoided by considering non-pharmacological treatments where available, e.g. CBT, good sleep hygiene and psychotherapy, especially in those not responding to pharmacotherapy.^24,25^ A meta-analysis of BZRAs in older adults for insomnia suggests that the magnitude of adverse effects may outweigh the clinical benefit received.^26^ There is also evidence that BZRAs could increase all-cause mortality risk,^27^ and dementia, especially when used for longer than three years.^28^ A number of interventions, involving patient education, tapering, pharmacological substitution, and psychological support, have been shown to be effective and feasible for deprescribing long-term BZRAs in older adults.^29^

The WHO’s Third Global Patient Safety Challenge, Medication without Harm, identifies transitions of care, polypharmacy, and vulnerable populations as priorities.^30^ Ensuring complete information is communicated when iniatiating medications in hospital is a means of targeting all of these priorities, and would change healthcare systems to support deprescribing, medication optimisation, and patient safety.

## Data Availability

Available on request.

## Acknowledgements

We acknowledge participating general practitioners and patients in the medication reconciliation study that provided data for these secondary analyses.

## References

1. Morgan K. Hypnotics in the elderly. Drugs. 1990;40(5):688–96.

2. Health at a Glance 2017 OECD Indicators [Internet]. 2017 [cited 8 August 2019]. Available from: https://www.oecd-ilibrary.org/docserver/health_glance-2017-en.pdf?expires=1565276694&id=id&accname=guest&checksum=544001399FE7FA4BF83B181F246388F8

3. Agarwal SD, Landon BE. Patterns in Outpatient Benzodiazepine Prescribing in the United States. JAMA Network Open. 2019;2(1):e187399–e187399.

4. Cadogan CA, Ryan C, Cahir C, Bradley CP, Bennett K. Benzodiazepine and Z-drug prescribing in Ireland: analysis of national prescribing trends from 2005 to 2015. British Journal of Clinical Pharmacology. 2018;84(6):1354–63.

5. Klotz U. Pharmacokinetics and drug metabolism in the elderly. Drug Metabolism Reviews. 2009;41(2):67–76.

6. O’Mahony D, O’Sullivan D, Byrne S, O’Connor MN, Ryan C, Gallagher P. STOPP/START criteria for potentially inappropriate prescribing in older people: version 2. Age and Ageing. 2015;44(2):213–8.

7. Fick DM, Semla TP, Beizer J, Brandt N, Dombrowski R, et al. American Geriatrics Society 2015 updated beers criteria for potentially inappropriate medication use in older adults. Journal of the American Geriatrics Society. 2015;63(11):2227–46

8. Berger A, Edelsberg J, Treglia M, Alvir JMJ, Oster G. Change in healthcare utilization and costs following initiation of benzodiazepine therapy for long-term treatment of generalized anxiety disorder: a retrospective cohort study. BMC Psychiatry. 2012;12(1):177.

9. Reeve E, To J, Hendrix I, Shakib S, Roberts MS, Wiese MD. Patient barriers to and enablers of deprescribing: a systematic review. Drugs & Aging. 2013;30(10):793–807.

10. Anderson K, Stowasser D, Freeman C, Scott I. Prescriber barriers and enablers to minimising potentially inappropriate medications in adults: a systematic review and thematic synthesis. BMJ Open. 2014;4(12):e006544.

11. Ahrens D, Chenot J-F, Behrens G, Grimmsmann T, Kochen MM. Appropriateness of treatment recommendations for PPI in hospital discharge letters. European Journal of Clinical Pharmacology. 2010;66(12):1265–1271.

12. Ahrens D, Behrens G, Himmel W, Kochen M, Chenot JF. Appropriateness of proton pump inhibitor recommendations at hospital discharge and continuation in primary care. International Journal of Clinical Practice. 2012;66(8):767–773.

13. Surendrakumar D, Dunn M, Roberts C. Hospital admission and the start of benzodiazepine use. BMJ: British Medical Journal. 1992;304(6831):881.

14. Stuffken R, van Hulten RP, Heerdink ER, Movig KL, Egberts AC. The impact of hospitalisation on the initiation and long-term use of benzodiazepines. European Journal of Clinical Pharmacology. 2005;61(4):291–295.

15. Grimmsmann T, Harden M, Fiß T, Himmel W. The influence of hospitalisation on the initiation, continuation and discontinuation of benzodiazepines and Z-drugs–an observational study. Swiss Medical Weekly. 2018;148:w14590.

16. Bell CM, Fischer HD, Gill SS, et al. Initiation of benzodiazepines in the elderly after hospitalization. Journal of General Internal Medicine. 2007;22(7):1024–1029.

17. Sinnott SJ, Bennett K, Cahir C. Pharmacoepidemiology resources in Ireland-an introduction to pharmacy claims data. European Journal of Clinical Pharmacology. 2017; 73(11): 1449–1455.

18. Soyombo S, Aujla H, Stanbrook R, et al. Socioeconomic deprivation and benzodiazepine/Z-drug prescribing: a cross-sectional study of practice-level data in England. British Journal of General Practice. 2019;69(suppl 1):bjgp19X703229.

19. Wilson S, Randell R, Galliers J, Woodward P. Reconceptualising clinical handover: information sharing for situation awareness. Paper presented at: ECCE 2009-European Conference on Cognitive Ergonomics 2009:258:315–322

20. Yemm R, Bhattacharya D, Wright D, Poland F. What constitutes a high quality discharge summary? A comparison between the views of secondary and primary care doctors. International Journal of Medical Education. 2014;5:125.

21. Wohlauer M. Fragmented care in the era of limited work hours: a plea for an explicit handover curriculum. BMJ Quality & Safety. 2012;21(Suppl 1):i16–i18.

22. Schwarz CM, Hoffmann M, Schwarz P, Kamolz L-P, Brunner G, Sendlhofer G. A systematic literature review and narrative synthesis on the risks of medical discharge letters for patients’ safety. BMC Health Services Research. 2019;19(1):158.

23. Newnham H, Barker A, Ritchie E, Hitchcock K, Gibbs H, Holton S. Discharge communication practices and healthcare provider and patient preferences, satisfaction and comprehension: a systematic review. International Journal for Quality in Health Care. 2017;29(6):752–768.

24. Guina J, Merrill B. Benzodiazepines I: Upping the care on downers: The evidence of risks, benefits and alternatives. Journal of Clinical Medicine. 2018;7(2):17.

25. Riemann D, Baglioni C, Bassetti C, et al. European guideline for the diagnosis and treatment of insomnia. Journal of Sleep Research. 2017;26(6):675–700. doi: 10.1111/jsr.12594.

26. Glass J, Lanctôt KL, Herrmann N, Sproule BA, Busto UE. Sedative hypnotics in older people with insomnia: meta-analysis of risks and benefits. BMJ. 2005;331(7526):1169.

27. Palmaro A, Dupouy J, Lapeyre-Mestre M. Benzodiazepines and risk of death: Results from two large cohort studies in France and UK. European Neuropsychopharmacology. 2015;25(10):1566–1577.

28. He Q, Chen X, Wu T, Li L, Fei X. Risk of Dementia in Long-Term Benzodiazepine Users: Evidence from a Meta-Analysis of Observational Studies. Journal of Clinical Neurology. 2019;15(1):9–19.

29. Reeve E, Ong M, Wu A, Jansen J, Petrovic M, Gnjidic D. A systematic review of interventions to deprescribe benzodiazepines and other hypnotics among older people. European Journal of Clinical Pharmacology. 2017;73(8):927–935. doi: 10.1007/s00228-017-2257-8.

30. Sheikh A, Dhingra-Kumar N, Kelley E, Kieny MP, Donaldson LJ. The third global patient safety challenge: tackling medication-related harm. Bulletin of the World Health Organisation. 2017;95(8):546–546A. doi: 10.2471/BLT.17.198002.

